# Factors associated with clinical severity in Emergency Department patients presenting with symptomatic SARS-CoV-2 infection

**DOI:** 10.1101/2020.12.08.20246017

**Authors:** Sophia Newton, Benjamin Zollinger, Jincong Freeman, Seamus Moran, Alexandra Helfand, Kayla Authelet, Matthew McHarg, Nataly Montano Vargas, Robert Shesser, Joanna Cohen, Derek Cummings, Yan Ma, Andrew C. Meltzer

**Affiliations:** The George Washington University, School of Medicine & Health Sciences, Department of Emergency Medicine. 2300 I Street, NW, Washington, DC 20037; George Washington University, Milken Institute School of Public Health, Department of Biostatistics and Bioinformatics. 950 New Hampshire Ave, NW, Washington, DC 20052; Children’s National Medical Center, Division of Emergency Medicine. 111 Michigan Ave NW, Washington, DC 20010; The George Washington University, School of Medicine & Health Sciences, Department of Pediatrics. 2300 I Street, NW, Washington, DC 20037; University of Florida, Department of Biology, Emerging Pathogens Institute. Bartram Hall, Gainesville, FL 32608

**Keywords:** Emergency Medicine, COVID-19 Disease, SARS-CoV-2 Infection, Infectious Diseases, Socioeconomic Factors

## Abstract

**Objective:** To measure the association of race, ethnicity, comorbidities, and insurance status with need for hospitalization of symptomatic Emergency Department (ED) patients with Severe Acute Respiratory Syndrome Coronavirus 2 (SARS-CoV-2) infection.

**Methods:** This study is a retrospective case-series of symptomatic patients presenting to a single ED with laboratory-confirmed SARS-CoV-2 infection from March 12-August 9, 2020. We collected patient-level information regarding demographics, public insurance status (Medicare or Medicaid), comorbidities, level of care, and mortality using a structured chart review. We compared demographics and comorbidities of patients who were (1) able to convalesce at home, (2) required admission to general medical service, (3) required admission to intensive care unit (ICU), or (4) died within 30 days of the index visit. Multivariable logistic regression analyses were performed to report adjusted odds ratios (aOR) and the associated 95% confidence intervals (95% CI) with hospital admission versus ED discharge home.

**Results:** In total, 993 patients who presented to the ED with symptoms were included in the analysis with 370 (37.3%) patients requiring hospital admission and 70 (7.1%) patients requiring ICU care. Patients requiring admission were more likely to be Black or African American, to be Hispanic or Latino, or to have public insurance (either Medicaid or Medicare.) On multivariable logistic regression analysis comparing which patients required hospital admission, African-American race (aOR 1.4, 95% CI 0.7-2.8) and Hispanic ethnicity (aOR 1.1, 95% CI 0.5-2.0) were not associated with need for admission but, public insurance (Medicaid: aOR 3.4, 95% CI 2.2-5.4; Medicare: aOR 2.6, 95% CI 1.2-5.3; Medicaid and Medicare: aOR 3.6 95% CI 2.1-6.2) and the presence of hypertension (aOR 1.8, 95% CI 1.2-2.7), diabetes (aOR 1.6, 95% CI 1.1-2.5), obesity (aOR 1.7, 95% CI 1.1-2.5), heart failure (aOR 3.9, 95% CI 1.4-11.2), and hyperlipidemia (aOR 1.8, 95% CI 1.2-2.9) were identified as independent predictors of hospital admission.

**Conclusion:** Comorbidities and public insurance are predictors of more severe illness for patients with SARS-CoV-2. This study suggests that the disparities in severity seen in COVID-19 among African Americans and Hispanics are likely to be closely related to low socioeconomic status and chronic health conditions and do not reflect an independent predisposition to disease severity.

## INTRODUCTION

Severe Acute Respiratory Syndrome Coronavirus 2 (SARS-CoV-2) was first reported in the United States in January 2020. Since then, there have been 7.5 million cases of SARS-CoV-2 infection with over 210,000 deaths in the US and rising.^1^ The clinical course of coronavirus disease 2019 (COVID-19), the disease caused by SARS-CoV-2, ranges widely in severity from asymptomatic to life-threatening. The mechanism behind this wide variation in patient outcomes is not yet well understood. Many studies thus far have documented associations between underlying patient comorbidities and more severe disease prognoses.^2-5^ In addition, reports suggest that Black and Hispanic populations are disproportionately impacted by COVID-19 disease with increased rates of infection compared to white populations.^6-8^ Understanding the reason for disparities among different patient populations is critical for targeting interventions aimed at mitigating the impact of disease. The objective of this study was to evaluate race, ethnicity, comorbidities, and insurance status as predictors of illness severity in patients presenting to the ED with symptomatic SARS-CoV-2 infection.

## METHODS

### Study Design and Setting

This was a single-center case series of patients with symptomatic SARS-CoV-2 infection presenting to an urban academic Emergency Department (ED) with approximately 70,000 annual visits. The Institutional Review Board at The George Washington University in Washington, DC approved the study with a waiver of consent on March 30, 2020.

### Study Population

Patients were included in this analysis if they presented to the ED with a chief complaint(s) consistent with COVID-19 and they tested positive for SARS-CoV-2 by nasopharyngeal swab using a polymerase chain reaction (PCR) platform. Initially all assays were performed at either the local Department of Health or commercial lab facility and subsequently tests were performed on one of the hospital’s testing platforms. Patients with positive SARS-CoV-2 results who were tested by protocol for another condition unrelated to COVID-19 such as trauma, intoxication, poisoning, suicidality, involuntary commitment, or isolated complaints highly unlikely to be related to COVID-19 (e.g. suture removal) were not included in this analysis. Additionally, asymptomatic, swab positive patients tested for reasons other than a clinician’s suspicion of COVID-19 disease were not included in this study. The decision to discharge, admit to a general medical floor, or admit to an ICU was made by the judgement of individual clinicians and typically based on a patient’s oxygen requirement and nursing needs.

### Data Collection

Patients were identified via monthly electronic health record (EHR) queries for positive tests for COVID-19 in the ED from March 12, 2020 until August 9, 2020. Data abstraction was performed after the index visit to capture 30-day outcomes. Chart review was performed according to guidelines outlined by Gilbert et al.^9^ Case report forms (CRFs) were created by Indiana University for use in a multi-center registry. All data was entered into a REDCap database. All data abstractors were trained specifically for this study and ten percent of charts were verified by a second abstractor for accuracy.

## Statistical Analysis

Data was initially stratified to identify significant associations by three categories of disposition: convalesce at home, admit to general medical service, and admit to intensive care unit (ICU) care. Patients who were initially discharged but then returned to the ED and needed admission were classified in the hospital admission category. Patients who were first admitted to the general wards but ultimately were transferred to ICU were included in the ICU category. Descriptive statistics were used to describe the demographics and comorbidities of patients per disposition. Frequencies and proportions were generated for categorical variables. Continuous variables were described using means and standard deviations. Methods for bivariate analyses were Chi-square or Fisher’s exact test for categorical data and analysis of variance (ANOVA) for continuous data.

Prior to regression analysis, variables were selected based on clinical knowledge and p-values <0.1 from bivariate analyses. We ran two separate logistic regression models: hospital admission vs. ED discharge and ICU care vs. general medical service admission. Independent variables included age, sex, race, ethnicity, health insurance, and comorbidities with need for hospitalization. In both models, patients who died within 30 days of the index visit were included in the more severe category (i.e. either needing hospital admission or ICU care). Adjusted odds ratios (aOR) and the associated 95% confidence intervals (95% CI) were reported; p-values <0.05 were considered statistically significant. All analyses were completed using SAS 9.4 (SAS Institute, Cary, NC).

## RESULTS

993 ED patients with viral syndromes and positive SARS-CoV-2 PCR assays were treated in the ED between March 12-August 9, 2020. Of these, 553 (55.7%) were discharged from the ED 370 (37.2%) were admitted to the floor, and 70 (7.0%) were admitted to the ICU. Descriptive statistics of patients were performed for all three categories (Table 1). Characteristics with statistically significant differences in disposition were age, race, ethnicity, and health insurance. In addition, there was statistically significant variation in disposition for multiple comorbid conditions: diabetes mellitus, hypertension, ischemic heart disease, obesity, hyperlipidemia, heart failure, atrial fibrillation, cancer, chronic obstructive pulmonary disease (COPD), and non-specified lung disease. When analyzed by logistic regression, age and health insurance were independent predictors of hospital admission (Table 2). Patients above the age of 40 were more likely to be admitted (40-64: aOR 2.1, 95% CI 1.4-3.4; ≥65: aOR 5.1, 95% CI 2.7-9.5) compared to those under 40. For patients with public health insurance, the odds ratio of being admitted were also significantly higher (Medicaid: aOR 3.4, 95% CI 2.2-5.4; Medicare: aOR 2.6, 95% CI 1.2-5.3; Medicaid and Medicare: aOR 3.6, 95% CI 2.1-6.2) than that of patients with private/commercial health insurance. Notably, neither race nor ethnicity were independent predictors of admission (Table 2). Of the comorbid conditions analyzed, diabetes mellitus, hypertension, obesity, hyperlipidemia, and heart failure were predictors of hospital admission (Table 2). Odds of admission or death were 1.6 (95% CI 1.1-2.5) times higher for patients with diabetes mellitus, 1.8 (95% CI 1.2-2.7) times higher for patients with systemic hypertension, 1.7 (95% CI 1.1-2.5) times higher for obese patients, 1.8 (95% CI 1.2-2.9) times higher for patients with hyperlipidemias, and 3.90 (95% CI 1.4-11.2) times higher for patients with heart failure when compared to previously healthy patients. When only admitted patients were analyzed to determine predictors of need for ICU care, patients with Medicaid were 2.2 times (95% CI 0.9-5.11) more likely to need ICU care compared to patients with private or commercial insurance (Table 3). A comorbid diagnosis of COPD also predicted need for ICU admission versus general medical service admission (aOR 2.5, 95% CI 1.01-6.1).

**Table 1.**
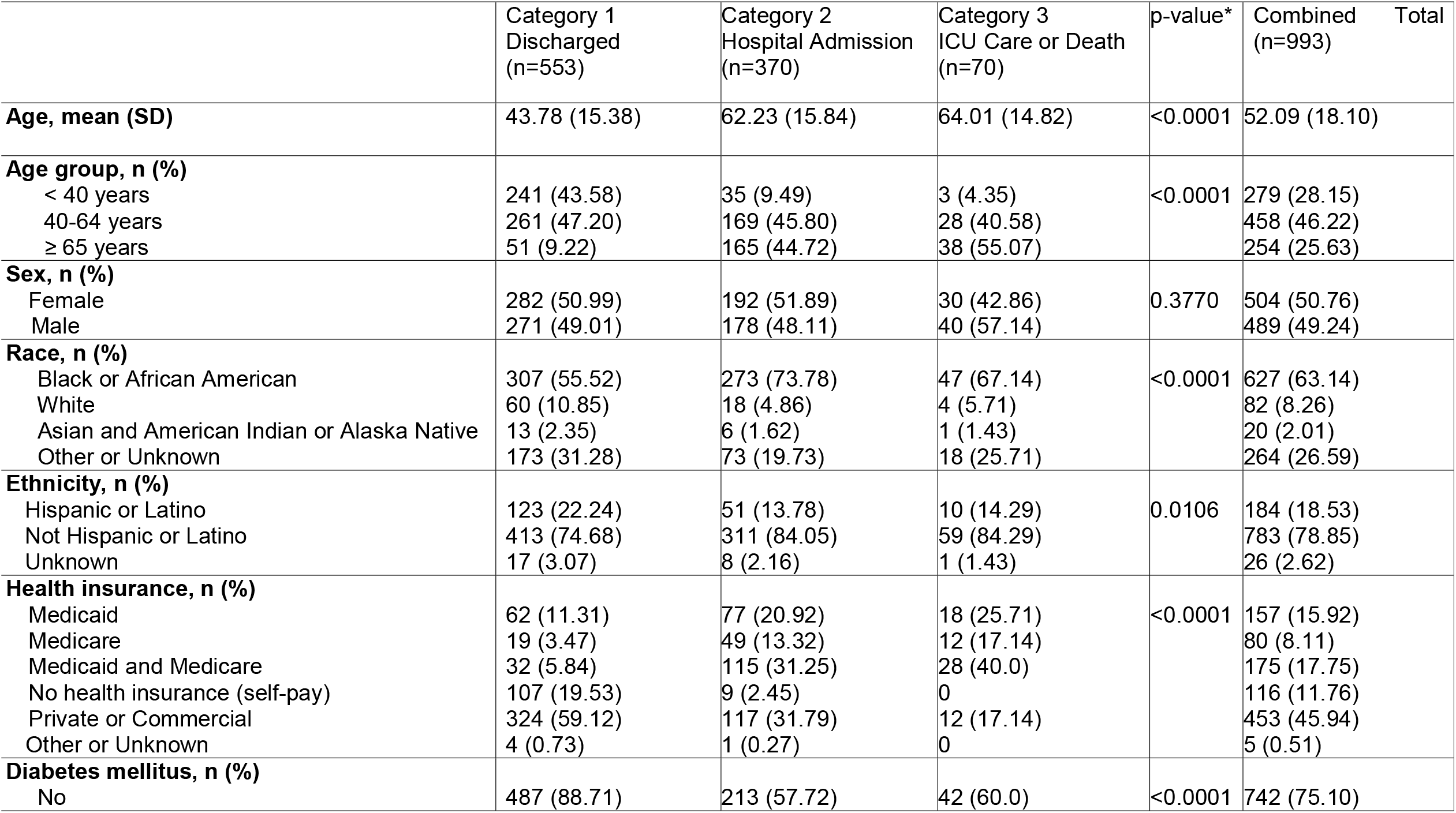
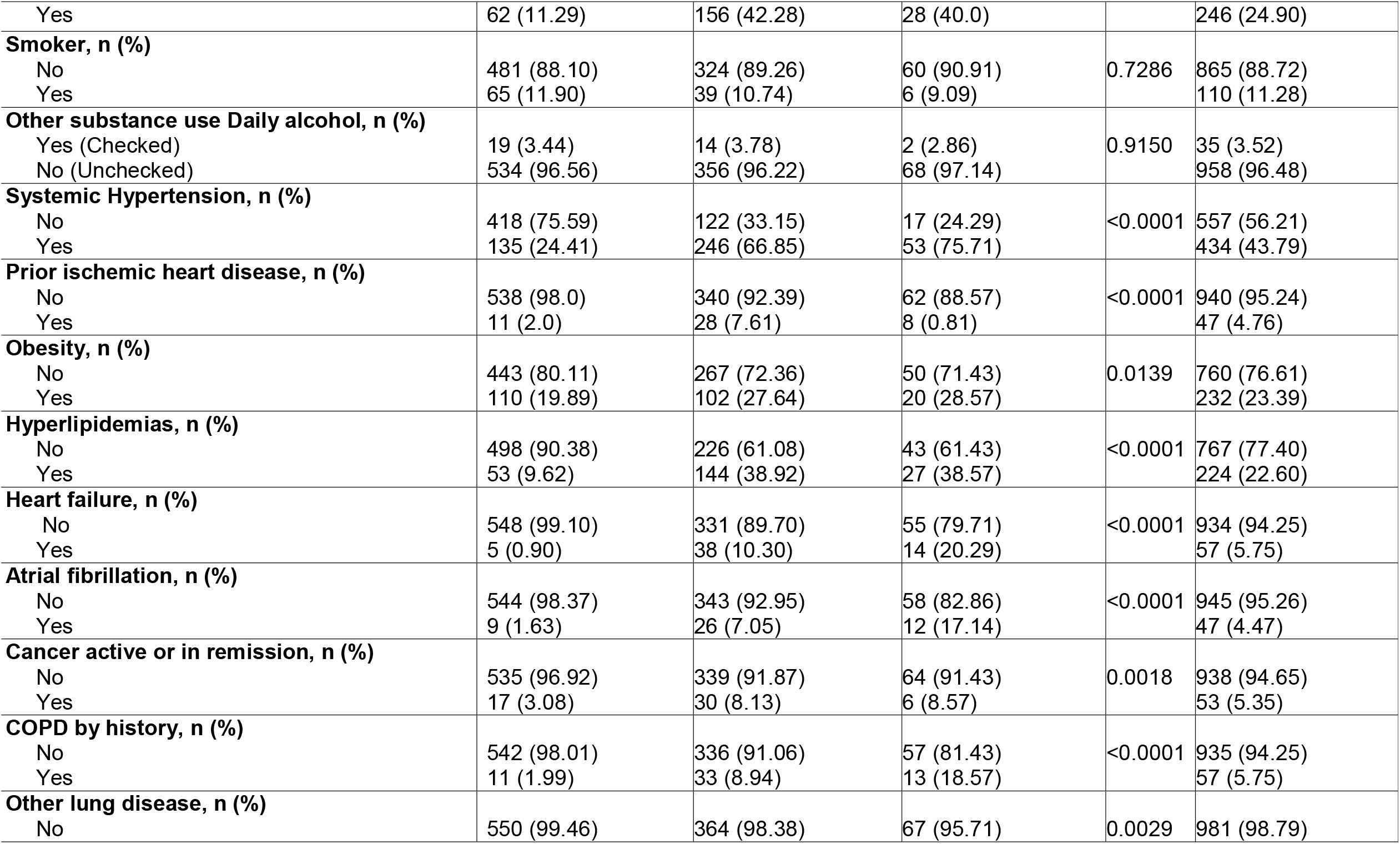
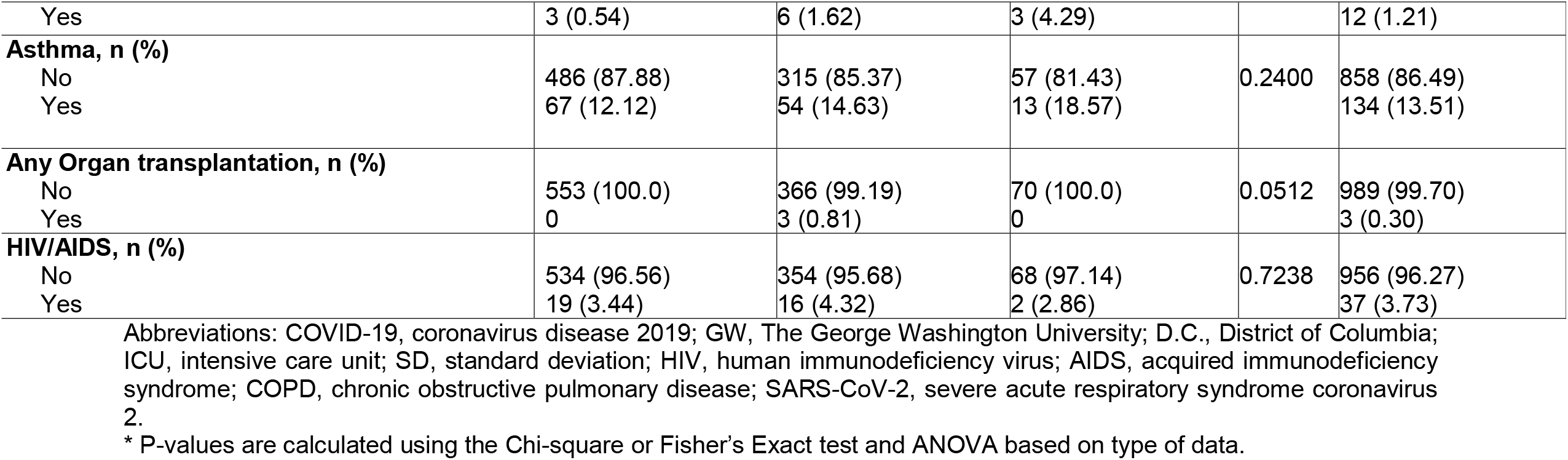
Characteristics of COVID-19 patients who presented at the GW Hospital Emergency Department in Washington.

**Table 2:**
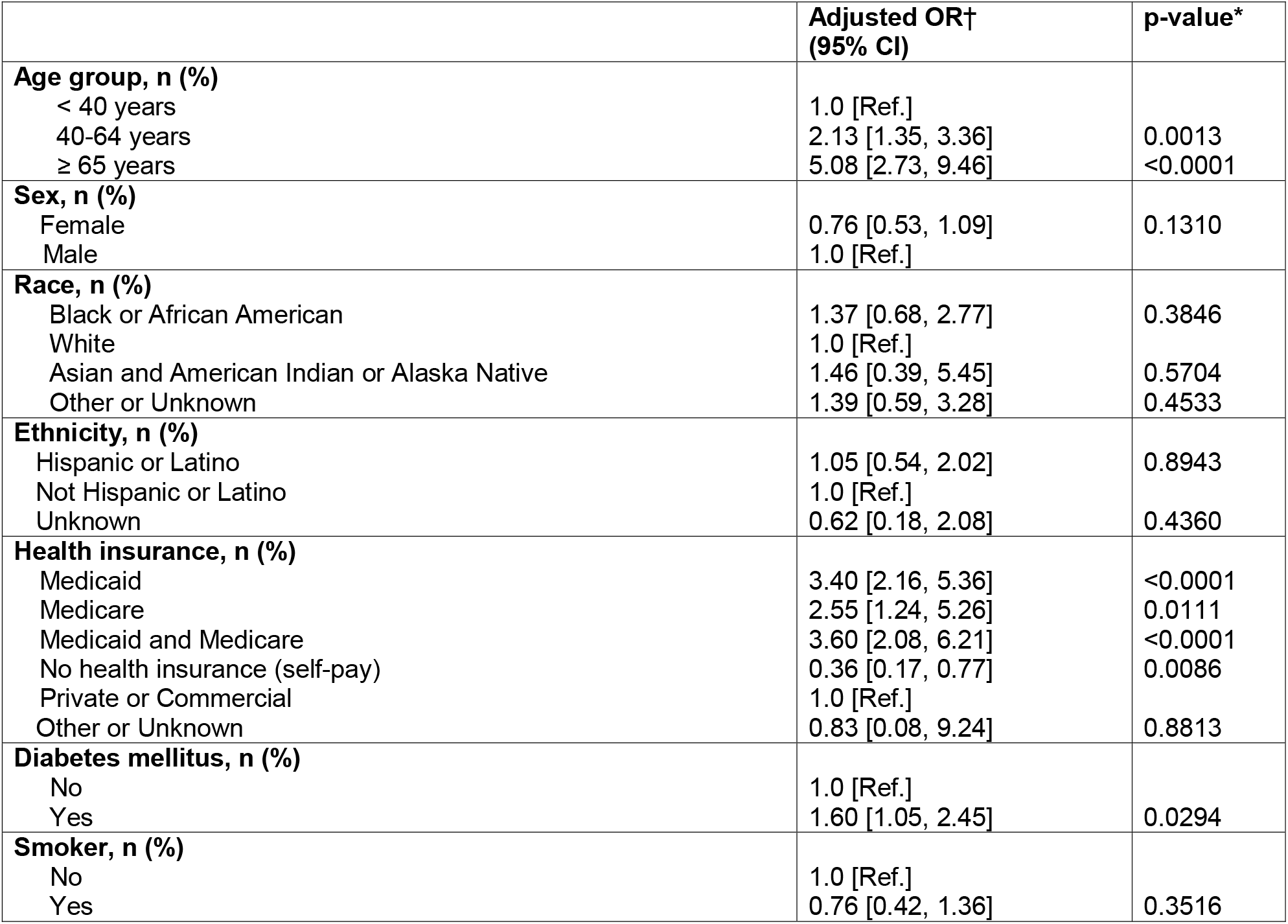
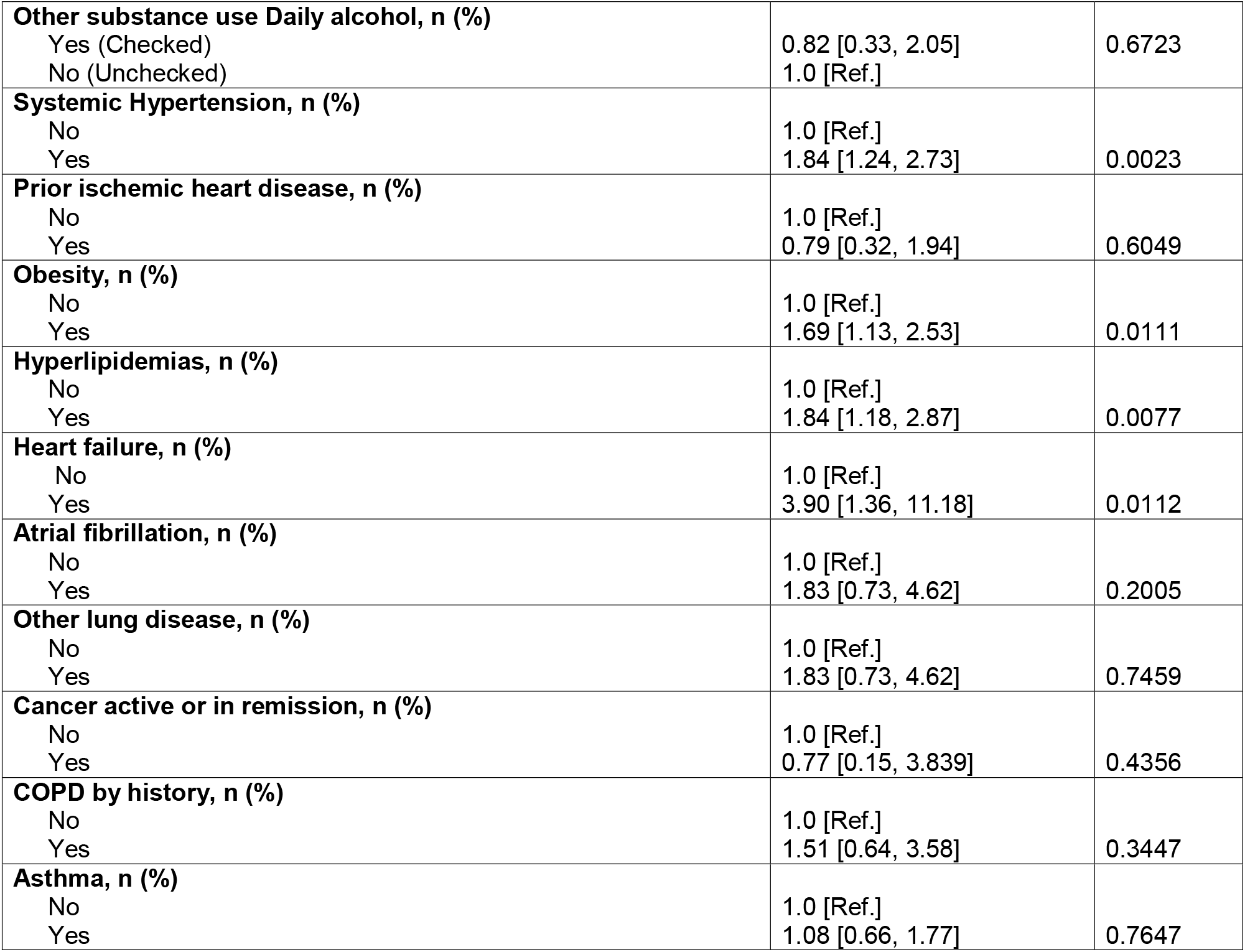
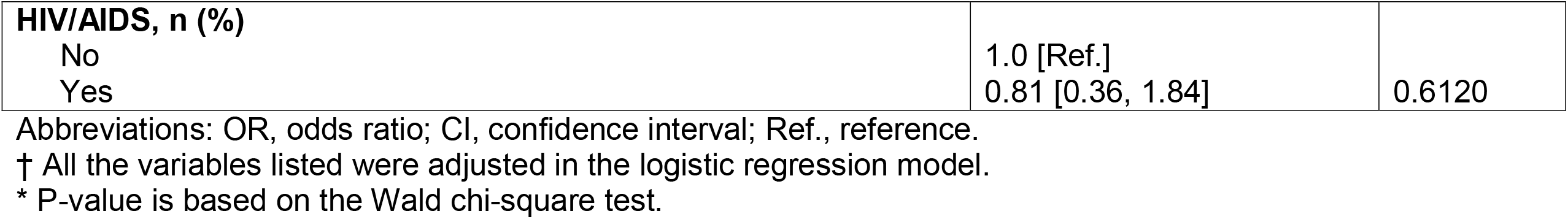
Logistic Regression Model for Hospital Admission or Death vs. Discharge.

**Table 3.** Logistic Regression Model for ICU care or Death vs. General Ward Admission.

|  | **Adjusted OR†**<br>**(95% CI)** | **p-value\*** |
| --- | --- | --- |
| **Age group, n (%)** |  |  |
| < 40 years | 1.0 [Ref.] |  |
| 40-64 years | 2.21 [0.47, 10.48] | 0.3193 |
| ≥ 65 years | 2.37 [0.46, 12.25] | 0.3040 |
| **Sex, n (%)** |  |  |
| Female | 0.67 [0.36, 1.26] | 0.2147 |
| Male | 1.0 [Ref.] |  |
| **Race, n (%)** |  |  |
| Black or African American | 0.62 [0.16, 2.37] | 0.4865 |
| White | 1.0 [Ref.] |  |
| Asian and American Indian or Alaska Native | 0.76 [0.05, 10.61] | 0.8360 |
| Other or Unknown | 1.37 [0.28, 6.62] | 0.6973 |
| **Ethnicity, n (%)** |  |  |
| Hispanic or Latino | 0.82 [0.25, 2.66] | 0.7392 |
| Not Hispanic or Latino | 1.0 [Ref.] |  |
| Unknown | 0.56 [0.06, 5.54] | 0.6181 |
| **Health insurance, n (%)** |  |  |
| Medicaid | 2.23 [0.97, 5.14] | 0.0607 |
| Medicare | 1.96 [0.67, 5.75] | 0.2231 |
| Medicaid and Medicare | 1.70 [0.70, 4.10] | 0.2414 |
| Private or Commercial | 1.0 [Ref.] |  |
| **Diabetes mellitus, n (%)** |  |  |
| No | 1.0 [Ref.] |  |
| Yes | 0.60 [0.32, 1.13] | 0.1157 |
| **Smoker, n (%)** |  |  |
| No | 1.0 [Ref.] |  |
| Yes | 0.54 [0.17, 1.68] | 0.2878 |
| **Other substance use Daily alcohol, n (%)** |  |  |
| Yes (Checked) | 0.38 [0.04, 3.64] |  |
| No (Unchecked) | 1.0 [Ref.] | 0.4006 |
| **Systemic Hypertension, n (%)** |  |  |
| No | 1.0 [Ref.] |  |
| Yes | 1.89 [0.89, 4.02] | 0.0975 |
| **Prior ischemic heart disease, n (%)** |  |  |
| No | 1.0 [Ref.] |  |
| Yes | 1.12 [0.42, 3.0] | 0.8256 |
| **Obesity, n (%)** |  |  |
| No | 1.0 [Ref.] |  |
| Yes | 1.35 [0.66, 2.78] | 0.4136 |
| **Hyperlipidemias, n (%)** |  |  |
| No | 1.0 [Ref.] |  |
| Yes | 0.74 [0.39, 1.41] | 0.3625 |
| **Heart failure, n (%)** |  |  |
| No | 1.0 [Ref.] |  |
| Yes | 1.74 [0.76, 3.96] | 0.1893 |
| **Atrial fibrillation, n (%)** |  |  |
| No | 1.0 [Ref.] |  |
| Yes | 1.62 [0.67, 3.94] | 0.2854 |
| **Other lung disease, n (%)** |  |  |
| No | 1.0 [Ref.] |  |
| Yes | 1.07 [0.18, 6.40] | 0.9445 |
| **Cancer active or in remission, n (%)** |  |  |
| No | 1.0 [Ref.] |  |
| Yes | 0.95 [0.33, 2.74] | 0.9220 |
| **COPD by history, n (%)** |  |  |
| No | 1.0 [Ref.] |  |
| Yes | 2.49 [1.01, 6.13] | 0.0468 |
| **Asthma, n (%)** |  |  |
| No | 1.0 [Ref.] |  |
| Yes | 1.11 [0.49, 2.51] | 0.7976 |
| **HIV/AIDS, n (%)** |  |  |
| No | 1.0 [Ref.] |  |
| Yes | 0.72 [0.13, 4.0] | 0.7072 |

Abbreviations: ICU, intensive care unit; OR, odds ratio; CI, confidence interval; Ref., reference.
† All the variables listed were adjusted in the logistic regression model.
\* P-value is based on the Wald chi-square test.
‡ No health insurance (self-pay) and Other or Unknown were excluded from the analysis due to 0 record.

## DISCUSSION

In this case-series of 993 patients presenting to an ED with symptomatic COVID-19, we identified both comorbidities and public insurance as predictors of illness severity. A history of hypertension, diabetes, heart failure, and hyperlipidemia were significant predictors disease severity. While Black patients were disproportionately represented in the cohort of patients requiring hospital admission or ICU care, this association was no longer evident after adjusting for insurance status and comorbidities.

Prior literature has shown that Black patients may have more severe disease that whites with COVID-19.^6,10-13^ Some sources suggest a bio-racial differential in immune response as the impetus for this disparity.^10^ When we adjusted our analysis for multiple factors including age, sex, ethnicity, health insurance, and comorbidities, the severity of illness in Black patients was not significantly different than White patients. Our study suggests that Black patients are at higher risk for severe COVID-19 outcomes due to the disproportionate burden of social determinates of health and chronic diseases such as hypertension, diabetes, and hyperlipidemia.^14,15^

In our population, public insurance, specifically Medicare or Medicaid, was an independent risk factor for severe COVID-19. The association between insurance type and severe COVID-19 persisted even when controlling for comorbidities. There are several possible reasons for this association. Firstly, it is possible that patients on public insurance may have more poorly controlled chronic conditions compared to patients with private insurance. Second, patients who experience poverty, and therefore are more likely to rely on Medicaid public insurance, may present later to the ED than those with private insurance in part due to decreased accessibility to outpatient healthcare.^16-18^

Third, public insurance may be associated with other unmeasured social determinants of health related to concerns about lost employment time, need for childcare, increased distance from the ED, and lack of transportation.^19-22^

When comparing the two models, we identified multiple independent risk factors for predicting need for admission versus discharge that were not identified when predicting need for general hospital admission versus ICU care. It is possible that patients who are sick enough to meet the threshold for general hospital admission (moderate severity) have similar risk factors to the patients who meet the higher threshold of ICU care (high severity). It is also possible that if the sample size for admitted patients was larger, we would see similar predictors for need for ICU care as we saw for hospital admission.

The actual mechanisms behind why certain comorbidities are associated with more severe outcomes are not yet well-understood. Blood sugar elevations have been proposed to play a role in weakening the immune response.^23-26^ The mechanistic relationship between uncontrolled hypertension and worse COVID-19 prognoses is not defined but hypertension has been associated with an increase in inflammatory markers and may be related to dysregulation of the renin-angiotensin-aldosterone axis.^27-31^ The severity of COVID-19 is likely the result of a complex interaction between viral load, host sex, host age, host comorbidities, social determinants, and host and viral genetic factors. The higher rates of COVID-19 in African-American and Latino populations has raised the suggestion that genetic susceptibility may play a role in severity. There have been limited studies suggesting a genetic predisposition to disease severity. One case report described SARS-CoV-2 deaths in three previously healthy adult brothers suggesting a genetic predisposition due to familial clustering.^32^ A second case report of SARS-CoV-2 of a large family cluster with more severe disease compared to other patients presenting at the same time also suggested a genetic predisposition due to apparent familial clustering of severity.^33^ Further research is needed to describe the role that host genetic factors play a role in disease susceptibility.^34^

### Strengths /Limitations

One strength of our study is the diverse characteristics of the patient population in terms of age, race, ethnicity, and health insurance status. In addition, we only included patients with symptoms of a viral-like illness and a laboratory-confirmed positive PCR test. Moreover, the dataset was relatively complete with few missing elements. Finally, we have high confidence in the data abstraction methods and utilized easily abstracted variables in order to remove ambiguity or subjectivity present in typical chart abstraction. This study has several important limitations. First, this study was reliant on documentation in the EHR of a single ED. In addition, we did not collect detailed socioeconomic data such as household income, educational attainment, and presence of primary care. Finally, we were unable to capture the severity and treatment status of comorbid conditions from chart review and, therefore, our results may not reflect the impact of underlying health conditions on the severity of COVID-19 for individual patients.

## Conclusion

Predictors of disease severity for ED patients who present with COVID-19 include age, Medicaid insurance, Medicare insurance, diabetes mellitus, hypertension, obesity, hyperlipidemia, and heart failure. Greater understanding of the factors that contribute to clinical variability in COVID-19 severity will assist in early identification of high-risk patients and enhance the precision of public health interventions.

## Data Availability

Data is currently at the lead site, Indiana University.

## References

1. CDC COVID data tracker. Centers for Disease Control & Prevention Web site. https://covid.cdc.gov/covid-data-tracker/#cases. Updated 2020.

2. Argenziano MG, Bruce SL, Slater CL, et al. Characterization and clinical course of 1000 patients with coronavirus disease 2019 in New York: retrospective case series. BMJ. 2020;369:m1996. Published 2020 May 29. doi:10.1136/bmj.m1996

3. Suleyman G, Fadel RA, Malette KM, et al. Clinical Characteristics and Morbidity Associated With Coronavirus Disease 2019 in a Series of Patients in Metropolitan Detroit. JAMA Netw Open. 2020;3(6):e2012270. Published 2020 Jun 1. doi:10.1001/jamanetworkopen.2020.12270

4. Targher G, Mantovani A, Wang XB, et al. Patients with diabetes are at higher risk for severe illness from COVID-19. Diabetes Metab. 2020;46(4):335–337. doi:10.1016/j.diabet.2020.05.001

5. Ebinger JE, Achamallah N, Ji H, et al. Pre-existing traits associated with Covid-19 illness severity. PLoS One. 2020;15(7):e0236240. Published 2020 Jul 23. doi:10.1371/journal.pone.0236240

6. Vahidy FS, Nicolas JC, Meeks JR, et al. Racial and ethnic disparities in SARS-CoV-2 pandemic: analysis of a COVID-19 observational registry for a diverse US metropolitan population. BMJ Open. 2020;10(8):e039849. Published 2020 Aug 11. doi:10.1136/bmjopen-2020-039849

7. Meyer P, Yoon P, Kaufmann R. CDC health disparities and inequalities report — united states, 2013. Centers for Disease Control & Prevention. 2013

8. Myers HF. Ethnicity-and socio-economic status-related stresses in context: an integrative review and conceptual model. J Behav Med. 2009;32(1):9–19. doi:10.1007/s10865-008-9181-4

9. Gilbert EH, Lowenstein SR, Koziol-McLain J, Barta DC, Steiner J. Chart reviews in emergency medicine research: Where are the methods?. Ann Emerg Med. 1996;27(3):305–308. doi:10.1016/s0196-0644(96)70264-0

10. Price-Haywood EG, Burton J, Fort D, Seoane L. Hospitalization and Mortality among Black Patients and White Patients with Covid-19. N Engl J Med. 2020;382(26):2534–2543. doi:10.1056/NEJMsa2011686

11. Yehia BR, Winegar A, Fogel R, et al. Association of Race With Mortality Among Patients Hospitalized With Coronavirus Disease 2019 (COVID-19) at 92 US Hospitals. JAMA Netw Open. 2020;3(8):e2018039. Published 2020 Aug 3. doi:10.1001/jamanetworkopen.2020.18039

12. Selden TM, Berdahl TA. COVID-19 And Racial/Ethnic Disparities In Health Risk, Employment, And Household Composition. Health Aff (Millwood). 2020;39(9):1624–1632. doi:10.1377/hlthaff.2020.00897

13. Assessment of COVID-19 hospitalizations by race/ethnicity in 12 states. Medical Letter on the CDC & FDA. Sep 6, 2020:9

14. Deere BP, Ferdinand KC. Hypertension and race/ethnicity. Curr Opin Cardiol. 2020;35(4):342–350. doi:10.1097/HCO.0000000000000742

15. Conway BN, Han X, Munro HM, et al. The obesity epidemic and rising diabetes incidence in a low-income racially diverse southern US cohort. PLoS One. 2018;13(1):e0190993. Published 2018 Jan 11. doi:10.1371/journal.pone.0190993

16. Chou SC, Gondi S, Weiner SG, Schuur JD, Sommers BD. Medicaid Expansion Reduced Emergency Department Visits by Low-income Adults Due to Barriers to Outpatient Care. Med Care. 2020;58(6):511–518. doi:10.1097/MLR.0000000000001305

17. Elek P, Molnár T, Váradi B. The closer the better: does better access to outpatient care prevent hospitalization? Eur J Health Econ. 2019 Aug;20(6):801–817. doi: 10.1007/s10198-019-01043-4. Epub 2019 Mar 15. PMID: 30877400; PMCID: PMC6652173.

18. Medical expenditure panel survey. Agency for Healthcare Research & Quality Web site. https://www.meps.ahrq.gov/mepsweb/about_meps/faq_results.jsp?ChooseTopic=All+Categories&keyword=&Submit2=Search

19. Alsan M, Stantcheva S, Yang D, Cutler D. Disparities in Coronavirus 2019 Reported Incidence, Knowledge, and Behavior Among US Adults. JAMA Netw Open. 2020;3(6):e2012403. Published 2020 Jun 1. doi:10.1001/jamanetworkopen.2020.12403

20. Centers for Medicare & Medicaid Services. Determining eligibility for Medicaid. Medicaid.gov Web site. https://www.medicaid.gov/medicaid/eligibility/index.html.

21. Rubin R. Household Composition May Explain COVID-19 Racial/Ethnic Disparities. JAMA. 2020;324(8):732. doi:10.1001/jama.2020.14375

22. Auger KA, Shah SS, Richardson T, et al. Association Between Statewide School Closure and COVID-19 Incidence and Mortality in the US. JAMA. 2020;324(9):859–870. doi:10.1001/jama.2020.14348

23. Bhandari S, Rankawat G, Singh A, Gupta V, Kakkar S. Impact of glycemic control in diabetes mellitus on management of COVID-19 infection [Published online ahead of print, 2020 Sep 2]. Int J Diabetes Dev Ctries. 2020;1–6. doi:10.1007/s13410-020-00868-7

24. Wang S, Ma P, Zhang S, et al. Fasting blood glucose at admission is an independent predictor for 28-day mortality in patients with COVID-19 without previous diagnosis of diabetes: a multi-centre retrospective study. Diabetologia. 2020;63(10):2102–2111. doi:10.1007/s00125-020-05209-1

25. Targher G, Mantovani A, Wang XB, et al. Patients with diabetes are at higher risk for severe illness from COVID-19. Diabetes Metab. 2020;46(4):335–337. doi:10.1016/j.diabet.2020.05.001

26. Wang J, Meng W. COVID-19 and diabetes: the contributions of hyperglycemia [Published online ahead of print, 2020 Oct 1]. J Mol Cell Biol. 2020;mjaa054. doi:10.1093/jmcb/mjaa054

27. Chen R, Yang J, Gao X, et al. Influence of blood pressure control and application of renin-angiotensin-aldosterone system inhibitors on the outcomes in COVID-19 patients with hypertension [Published online ahead of print, 2020 Oct 2]. J Clin Hypertens (Greenwich). 2020;10.1111/jch.14038. doi:10.1111/jch.14038

28. Meng J, Xiao G, Zhang J, et al. Renin-angiotensin system inhibitors improve the clinical outcomes of COVID-19 patients with hypertension. Emerg Microbes Infect. 2020;9(1):757–760. doi:10.1080/22221751.2020.1746200

29. Samidurai A, Das A. Cardiovascular Complications Associated with COVID-19 and Potential Therapeutic Strategies. Int J Mol Sci. 2020;21(18):6790. Published 2020 Sep 16. doi:10.3390/ijms21186790

30. Rodilla E, Saura A, Jiménez I, et al. Association of Hypertension with All-Cause Mortality among Hospitalized Patients with COVID-19. J Clin Med. 2020;9(10):E3136. Published 2020 Sep 28. doi:10.3390/jcm9103136

31. F Fang L, Karakiulakis G, Roth M. Are patients with hypertension and diabetes mellitus at increased risk for COVID-19 infection? [Published correction appears in Lancet Respir Med. 2020 Jun;8(6):e54]. Lancet Respir Med. 2020;8(4):e21. doi:10.1016/S2213-2600(20)30116-8

32. Yousefzadegan S, Rezaei N. Case Report: Death due to COVID-19 in Three Brothers. Am J Trop Med Hyg. 2020;102(6):1203–1204. doi:10.4269/ajtmh.20-0240

33. Patarčić I, Gelemanović A, Kirin M, et al. The role of host genetic factors in respiratory tract infectious diseases: systematic review, meta-analyses and field synopsis. Sci Rep. 2015;5:16119. Published 2015 Nov 3. doi:10.1038/srep16119

34. LoPresti M, Beck DB, Duggal P, Cummings DAT, Solomon BD. The Role of Host Genetic Factors in Coronavirus Susceptibility: Review of Animal and Systematic Review of Human Literature. Am J Hum Genet. 2020;107(3):381–402. doi:10.1016/j.ajhg.2020.08.007

